# Psychological impact of the coronavirus disease 2019 (COVID-19) outbreak on healthcare workers in China

**DOI:** 10.1101/2020.03.03.20030874

**Authors:** Yuhong Dai, Guangyuan Hu, Huihua Xiong, Hong Qiu, Xianglin Yuan

**Author notes:** **Corresponding Author:** Xianglin Yuan, M.D., Ph.D., Affiliation:1. Tongji Hospital,Tongji Medical College, Huazhong University of Science and Technology, 2. The Second Medical Clinical College, Tongji Medical College, Huazhong University of Science and Technology, University, Wuhan 4300730 Hubei, China, No.1095 Jie Fang Avenue, Hankou, Wuhan 430030, P.R. China., Hong Qiu, M.D., Tongji Hospital,Tongji Medical College, Huazhong University of Science and Technology, No.1095 Jie Fang Avenue, Hankou, Wuhan 430030, P.R. China. Contributed equally.

## Abstract

**Introduction:** Since the outbreak of coronavirus disease 2019 (COVID-19), more than 3000 (including clinical diagnosis) healthcare workers (HCWs) have been infected with severe acute respiratory syndrome coronavirus 2 (SARS-CoV-2) in China. This study is aimed to investigate the risk perception and immediate psychological state of HCWs in the early stage of the COVID-19 epidemic.

**Methods:** This study utilized a cross-sectional survey designed on a convenient sample of 4357 HCWs in China. Its data were collected using anonymous structured questionnaires distributed through social software. 6 questions were set to evaluate the participants’ risk perception of COVID-19, and a General Health Questionnaire was used to identify the participants’ immediate psychological status. Descriptive statistics were used for data analysis. Risk perception and psychological status were compared by demographic characteristics and COVID-19 exposure experiences.

**Result:** A total of 4,600 questionnaires were distributed, and 4,357 qualified ones (94.7%) were collected. The main concerns of HCWs are: infection of colleagues (72.5%), infection of family members (63.9%), protective measures (52.3%) and medical violence (48.5%). And 39.1% of the HCWs had psychological distress, especially working in Wuhan, participating in frontline treatments, having been isolated and having family members or colleagues infected.

**Conclusions:** The finding indicating that, faced with the COVID-19 epidemic, HCWs, especially in Wuhan, were worried about the risks of infection and protective measures, resulting in psychological distress, so further actions should be taken.

## INTRODUCTION

Since December 2019, the outbreak of COVID-19 has developed in Wuhan, Hubei Province, China, surrounding the epicenter of Huanan Seafood Market, a local wet market. In the following weeks, the epidemic had spread rapidly, with the number of suspected and confirmed cases steadily rising, resulting in a magnitude of distribution far exceeding that of severe acute respiratory syndrome (SARS) in 2003. More than 60,000 HCWs are participating in handling and treating the COVID-19 patients in Wuhan. COVID-19 is characterized by its unambiguous capacity of person-to-person transmission^1 2^, which was not clearly recognized initially. As a consequence, the protection for HCWs was sub-optimal, and occupational exposures and infections were frequent.^3^

As the number of cases climbed up at a great speed, panics prevailed among the general population and the capacity of local fever clinics was soon overwhelmed. The speedily increasing number of COVID-19 cases prevented some confirmed or suspected patients from receiving timely treatment. As a result, tensions and anxieties were further fueled between patients and HCWs, with violence against the latter occurring. By February 3, a total of 20,438 confirmed cases, 23,214 suspected cases and 425 deaths had been reported in 31 province-level administrative regions in China.^4^ And familial clusters are also observed.^2^ Under such a medical and social background, HCWs are facing a multitude of challenges: an abrupt outbreak of epidemic, a steep-ascending workload, a substantial risk of occupational exposure and violence, a high risk of infection of themselves and their family members, and insufficient supply of protective materials. The psychological response of overloaded HCWs is of great importance with regard to the effect of the defense against the epidemic. We therefore investigated the general condition of HCWs in the early phases of the COVID-19 outbreak in Wuhan as well as their perception of risks and psychological states.

## METHODS

### Study design and Participants

The study was a cross-sectional survey, a convenience sample of 4,357 HCWs from Wuhan, other regions of Hubei province, and other provinces was gathered. Starting on February 3, 2020, the study lasted for eight days, during which, the number of confirmed and suspected cases was increasing rapidly. As the epidemic can be transmitted through droplets and contacts, in order to avoid interpersonal contacts, structured questionnaires were forwarded online to all HCWs through social software, and the respondents could decide whether to participate in the survey. The study was approved by the Ethics Committee of Tongji Hospital of Tongji Medical College of Huazhong University of Science and Technology, and was registered with ClinicalTrials.gov, number NCT04260308. Only a single response to the questionnaire was permitted for each person. The questionnaire consisted of four parts: basic demographic data, the occupational exposure experience, risk perception of COVID-19, and the General Health Questionnaire.

### Basic demographic data, Exposure to COVID-19 and Risk perception

The information of basic demography was collected, such as age, gender, marital status and work experience. And the COVID-19 exposure experience was also recorded in the survey. 6 questions were designed to investigate the participants’ perception of risk to COVID-19, which were assessed on a five-point Likert scale (1. strongly worried; 2. worried; 3. not sure; 4. not too worried; 5. not worried at all), with score points of 1 to 5 assigned, a lower score indicating a higher level of concern. The 6 questions are: 1. Are you worried about getting infected with COVID-19 yourself? 2. Are you worried about your family members getting infected with COVID-19 from you? 3.Are you worried about medical violence? 4. Are you worried about colleagues at the frontline (direct contact with the COVID-19 patients)? 5. Are you worried about inadequate protective measure? And 6. Are you worried about the current grassroots prevention and control strategy?

### GHQ-12

General Health Questionnaire is a widely accepted mental health measurement, with high reliability and validity. It has been translated into many languages and used in many countries. Moreover, GHQ-12 showed a high positive rate in the evaluation of people’s psychological state after the SARS epidemic ^5 6^ and Wenchuan earthquake^7 8^ in Chinese population. We adopted the scoring method of 0-0-1-1.^9^ During the study period, the workload of HCWs was huge. Take Wuhan as an example: on February 3, 2020, a total of 8,279 COVID-19 patients were admitted in 28 designated hospitals.^10^ In order not to increase the burden of HCWs, we chose only 12 questions to screen out the HCWs who may have psychological distress and need attention and intervention in a simple and convenient way.

### Statistical analysis

The data were analyzed by using the Statistical Product and Service Solutions 22.0 (SPSS 22.0). We first analyzed the exposure experience to COVID-19, and then compared different region groups by means of chi-square tests. Independent t-test and ANOVA test were used to analyze the five-point Likert scale. Finally, variables significant at the level 0.05 were included in multivariate analysis to identify the independent prognostic factors of psychological distress.

### Patient and Public Involvement statement

Our study was carried out on HCWs, PPI representatives worked with us to refine the research question. We have invited HCWs participated in this study to help us develop our dissemination strategy.

## RESULT

### Demographic data and COVID-19 exposure experience

A total of 4,600 questionnaires were distributed, and 4,357 qualified ones (94.7%) were collected. Of the respondents, 1,026 (23.5%) were males and 3,331 (76.8%) females; and 1,419 (32.5%) doctors while 2,343 (53.8%) nurses. The mean age of the participants was 35 years old (standard deviation, SD:8.6, range: 20-81 years old). Demographic characteristics of the study’s respondents are listed in Table 1.

**Table 1.** Demographic characteristics of the study respondents

|  | N=4357(%) |
| --- | --- |
| Gender |  |
| male | 1026(23.5) |
| female | 3331(76.5) |
| Age |  |
| ≤25 years | 453(10.4) |
| 26-40 years | 2901(66.6) |
| 41-60 years | 983(22.5) |
| > 60 years | 20(0.5) |
| Occupation |  |
| Doctor | 1419(32.6) |
| Nurse | 2343(53.8) |
| Technician | 437(10.0) |
| Support staff | 158(3.6) |
| Location |  |
| Wuhan | 1339(30.7) |
| Other regions in Hubei | 2364(54.3) |
| Other provinces | 654(15.0) |
| Working life |  |
| < 2 years | 389(8.9) |
| 2-5 years | 931(21.4) |
| 6-10 years | 1077(24.7) |
| > 10 years | 1960(45.0) |
| Hospital level |  |
| Ministerial/provincial | 1202(27.6) |
| Municipal | 1913(43.9) |
| Country | 734(16.8) |
| Township/Community | 508(11.7) |
| Marital status |  |
| Married | 3338(76.6) |
| Unmarried | 947(21.7) |
| Others | 72(1.7) |
| Fertility status |  |
| With children | 3087(70.9) |
| Childless | 1270(29.1) |

During the study period, 1,404 (32.2%) of the 4,357 HCWs were participating in the frontline treatment of COVID-19 patients. Among the 1,339 (30.7%) HCWs enrolled in Wuhan, 516 (38.5%) were participating in the frontline treatment, including 133 doctors (25.8%) and 326 nurses (63.2%). In this study, 40 HCWs were infected with COVID-19, 33 of whom were from Wuhan, 6 from other areas of Hubei province, and only one from other provinces. 89.9% of the respondents expressed their willingness to participated in the treatment. Compared with other regions, COVID-19 exposure was even more severe in Wuhan. COVID-19 exposure experiences differentiated by locations are depicted in Table 2.

**Table 2.** Exposure experience of COVID-19, by locations

| | Wuhan(n=1339) | Other regions in Hubei(n=2364) | Other provinces (n=654) | $\chi^2$ |
| --- | --- | --- | --- | --- |
|  | No (%) | No (%) | No (%) |  |
| Participate in treatment |  |  |  | 176.912* |
| Frontline | 516(38.5) | 738(31.2) | 150(22.9) |  |
| Upcoming | 245(18.3) | 227(9.6) | 44(6.7) |  |
| Possible | 404(30.2) | 986(41.7) | 307(46.9) |  |
| Impossible | 174(13.0) | 413(17.5) | 153(23.4) |  |
| Isolated |  |  |  | 54.492* |
| Yes | 270(20.2) | 284(12.0) | 70(10.7) |  |
| No | 1069(79.8) | 2080(88.0) | 584(89.3) |  |
| Self-infection |  |  |  | 50.880* |
| Yes | 33(2.5) | 6(0.3) | 1(0.2) |  |
| No | 1306(97.5) | 2358(99.7) | 653(99.8) |  |
| Family members infected |  |  |  | 139.769* |
| Yes | 137(10.2) | 59(2.5) | 6(0.9) |  |
| No | 1202(89.8) | 2305(97.5) | 648(99.1) |  |
| Colleagues infected |  |  |  | 902.971* |
| Yes | 781(58.3) | 434(18.4) | 24(3.7) |  |
| No | 558(41.7) | 1930(81.6) | 630(96.3) |  |
| Volunteers |  |  |  | 3.835 |
| Yes | 1221(91.2) | 2114(89.4) | 518(88.8) |  |
| No | 118(8.8) | 250(10.6) | 73(11.2) |  |

\* $p < 0.05$

### Risk perception

The main concerns of HCWs were: infection of colleagues (72.5%), infection of family members (63.9%), protective measures (52.3%) and medical violence (48.5%). Only 34.7% of HCWs expressed very worried about the risk of self-infection. About half of the respondents expressed confidence (46.3%) or high confidence (9.1%) in the current grassroots prevention and control strategy. The distribution of risk perception is shown in Figure 1.

**Figure 1.**
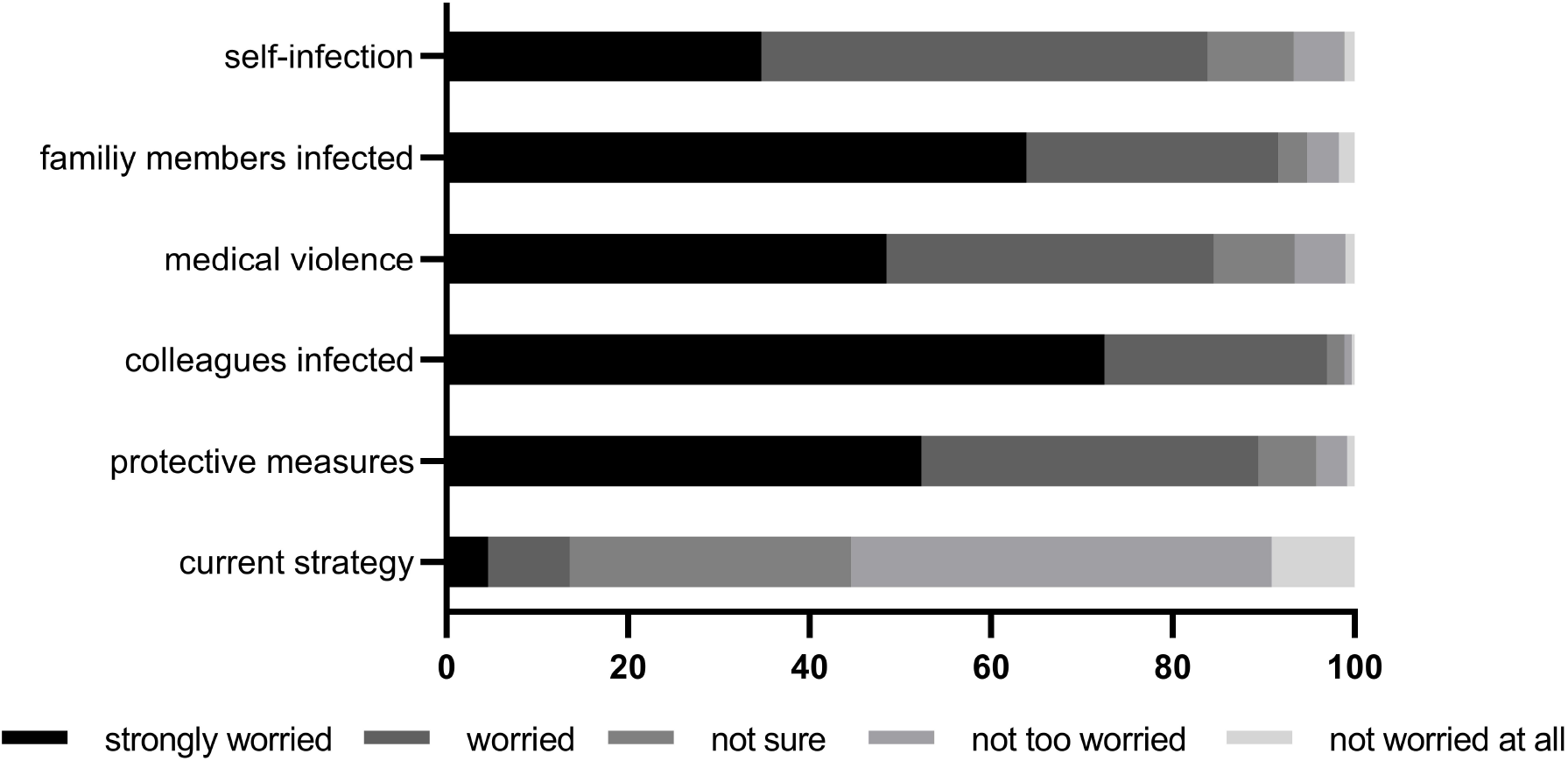
Distribution of risk perception to COVID-19.

However, 40% of frontline HCWs expressed very worried about getting infected, significantly higher than other three groups; and the score of Likert scales were much lower (F=49.55, *p*=0.000). We found that compared with other regions, all of the 6 questions had lower scores, showing significant statistical differences in Wuhan. And the HCWs in Wuhan had insufficient confidence in the current prevention and control strategy.

In different treatment division of labor, nurses were more worried about their own infection at work than doctors, technicians and support personnel (F = 33.262, *p*= 0.000).

Female respondents were more worried about infection and violence, and those who had children were more worried about family members’ infection (F = 59.472, *p*= 0.000) and medical violence (F = 3.056, *p*= 0.080).

### Psychological status

The scores of GHQ-12 for the general population were in a positive skew distribution, with a score range of 0-12 and a median of 2 (SD: 2.56). Taking the total score of 3 as the cut-off value, ≥ 3 points is considered a case that there may have psychological distress, which needs further confirmation and intervention. Of the 4,357 HCWs, 1,704 (39.1%) had psychological distress.

Through the analysis of the basic demography and COVID-19 exposure experience, the relevant factors affecting the mental health of the HCWs were confirmed. Through one-way ANOVA test, 8 demographic data (gender, age, location, occupation, work life, hospital level, marital status and fertility status) and 6 work exposure conditions (participating in the frontline treatment, having been isolated, getting infected, having family members infected, having colleagues infected, and volunteering for treatment) were all related to the psychological state. Through multiple factor regression analysis, it was found that women, working in Wuhan area and working in primary hospitals were poor prognostic factors. In terms of COVID-19 exposure history, poor prognostic factors include participating in frontline treatment, involuntarily participating in treatment work, having been isolated, and having family members or colleagues infected, while whether the respondents themselves have got infected or not was not an independent prognostic factor of psychological disorder. Risk factors for GHQ are depicted in Table 3.

**Table 3.**
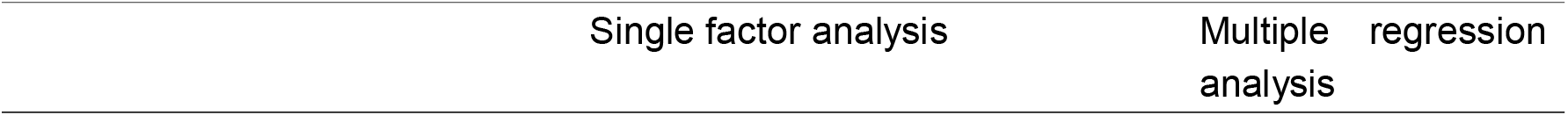
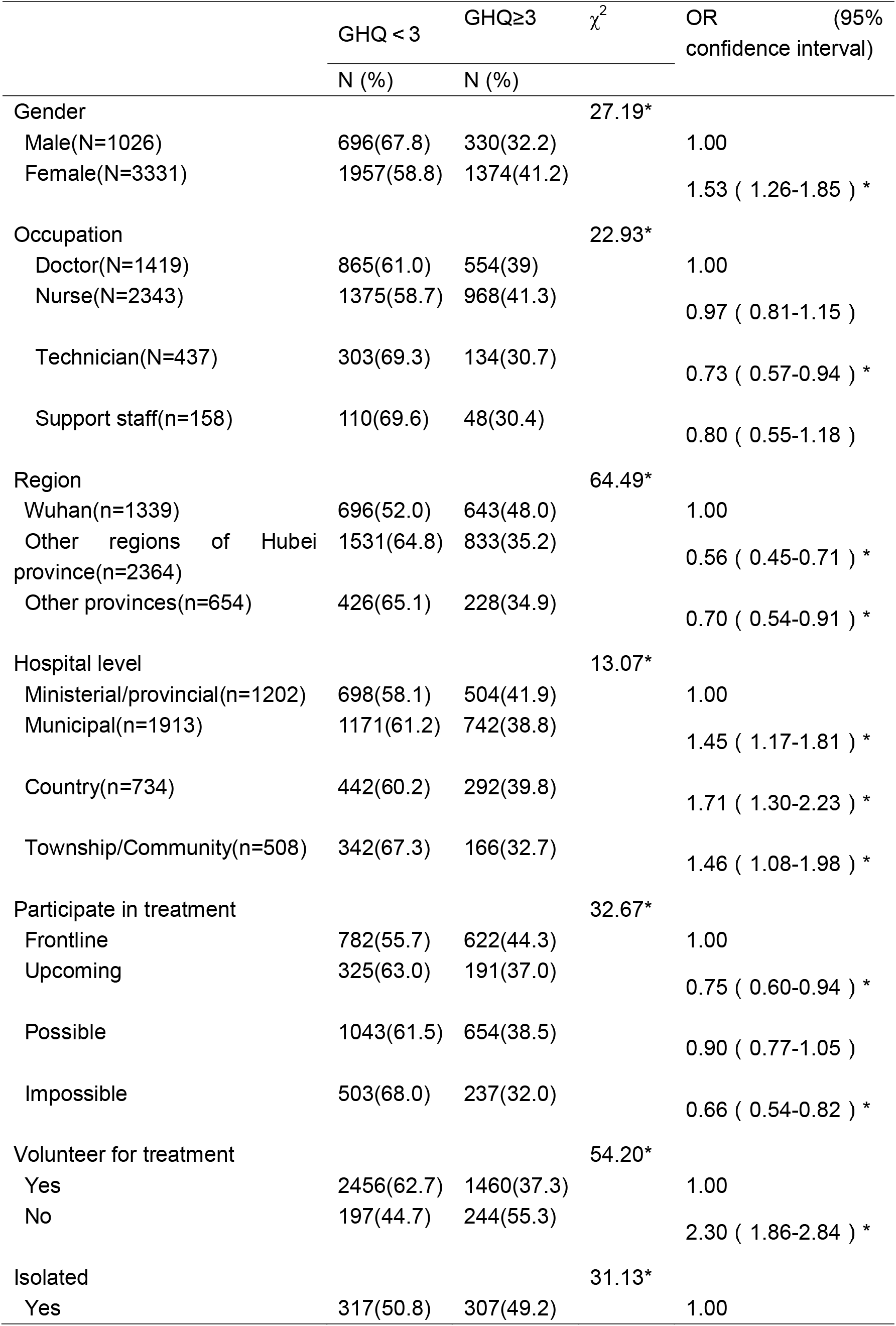
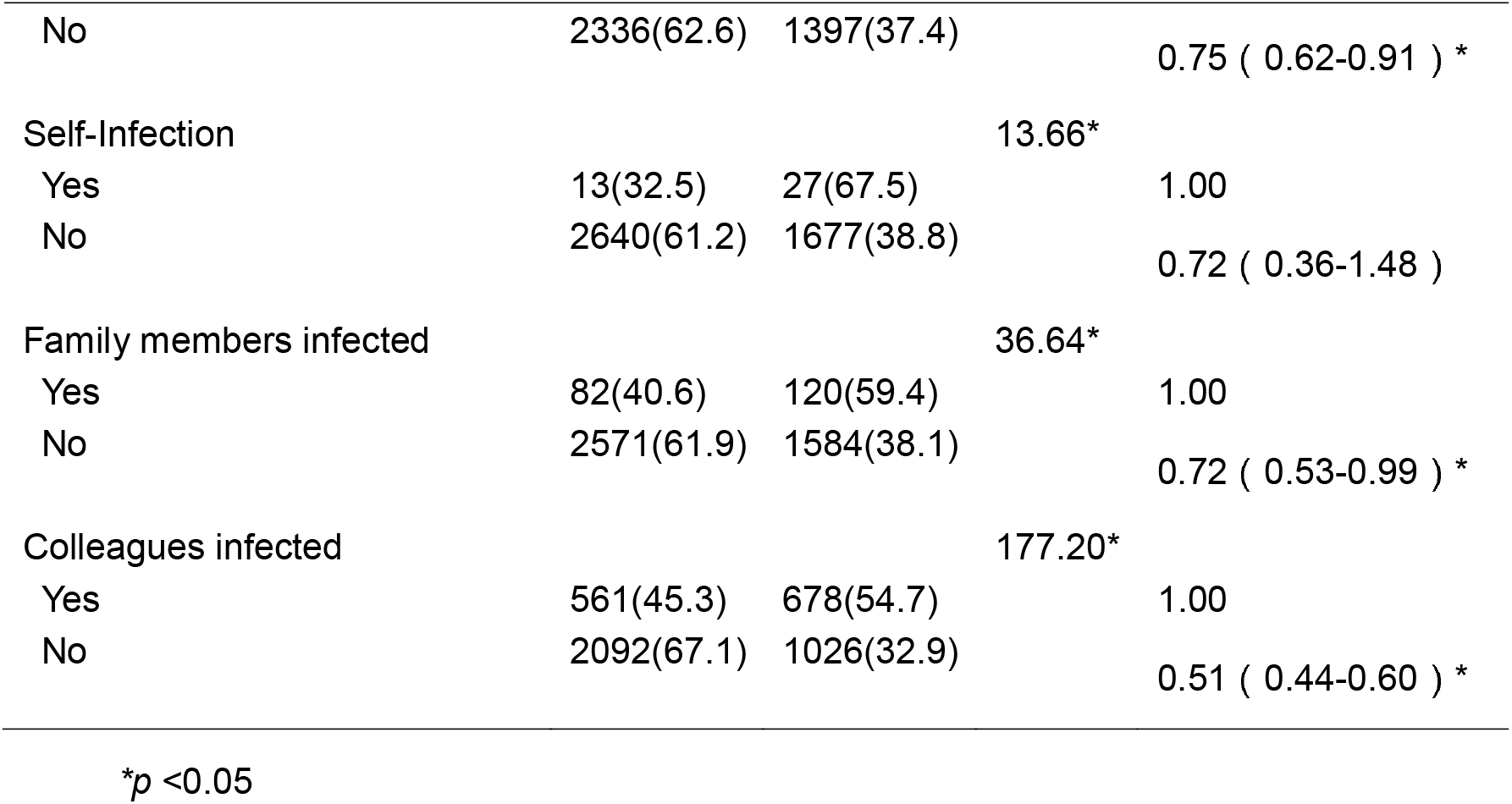
Regression analysis of GHQ-12 score

## DISCUSSION

Since mid-December 2019 when COVID-19 broke out in Wuhan, it has spread rapidly. Since January 23, 2020, the city blockade has been carried out in Wuhan, and traffic restrictions were enforced throughout China. People in Wuhan, even across China, live on the horizon of COVID-19. The pathogen of this disease has been identified as coronavirus, which has 86.9% nucleotide sequence homology with the bat SARS-like coronavirus genome.^11^ According to the official statistics, 44,745 cases had been diagnosed in China, and 398 in areas out of China, until the day this study ended, February 11, 2020. Its impressing ability of spreading contagion exceeds SARS in 2003.^12^

In the initial stage, this disease didn’t arouse enough attention from administration departments, and the Chinese traditional Spring Festival accelerated the spread of epidemic across the whole nation, because the Spring Festival is the largest population migration annually. The arrival of the Spring Festival cut the medical protection industry’s capacities, which further aggravated the phenomenon of insufficient protection. The rapidly uprising number of patients, and the public panics after it proved highly contagious, plus shortage of protective supplies and frequent occurrences of violent events in medical facilities ^13^, seemed to make medical burdens even heavier. 1,716 HCWs were infected and 6 staff died of this disease, according to the Chinese official announcement, which is thought to be incomplete statistics. We should pay more attention to HCWs’ psychological tolerance, especially those fighting at the frontline of controlling the plaque.

In this study, we used GHQ-12 to identify the mental health of HCWs. The criterion for determining suspected mental health problems is defined as a score of ≥3 points. The survey results of Chinese residents by using the GHQ-12 questionnaire showed that the positive rate of residents with a GHQ-12 score of ≥3 points is about 12.8%-21.18%.^14^ Many studies have used this scale to evaluate scores of HCWs in different countries, showing a positive rate of 25% −32%.^14-16^ A survey of frontline HCWs in Hong Kong during the period of SARS through GHQ-12 questionnaire showed 57% having experienced psychological distress.^17^ In this study, 39.1% of respondents were with GHQ-12 score ≥3, it is suggested that the incidence of psychological abnormality was significantly higher than that in normal times.

A higher proportion of HCWs from Wuhan showed psychological abnormalities. As Wuhan is the epidemic focus of COVID-19, there are a huge number of patients, high rate of severe illness and high mortality. Due to the shortage of HCWs in Wuhan, they have to work with high workload for a long time. In addition, 10.2% of HCWs in Wuhan reported that their relatives were infected, while only 2.5% and 0.9% in other regions of Hubei and other provinces, respectively. And in Wuhan, the COVID-19 infecting rate among HCWs is much higher than other regions, reaching 20.2%. 58.3% respondents in Wuhan indicated their colleagues were infected, while only 18.4% and 3.7% in other regions of Hubei and other provinces, respectively. In Wuhan, 38.5% of the HCWs directly participated in the frontline treatment of the COVID-19 patients. And through multiple regression analysis, we found that psychological abnormality rate was much higher in the frontline HCWs. It is easy to understand that the frontline HCWs needed to contact COVID-19 patients directly and might be equipped with insufficient protective measures at any time, so they show more concerns for their own safety. Similarly, nurses had longer periods of contacts with patients; and in the infectious departments, because of no caregivers allowed in, nurses needed to take care of all the life of patients, thus more worried about getting infected. However, although the proportion of nurses with psychological disorder was higher than that of other professions, there was no significant difference. In primary hospitals, the lack of protective measures was more obvious, and primary HCWs lacked experience in treating critical illness, so there was a higher rate of psychological distress.

Before and during the period of COVID-19 epidemic, incidents of violence against HCWs occurred in some parts of the country, including in Wuhan. In answering the questions on risk perception, 56.8% of the health workers said they were very worried about the medical violence, while 43.7% and 48.3% in other regions of Hubei and other provinces, respectively. How to prevent the medical violence and protect the HCWs in the upfront line of defense against COVID-19 is another critical problem for Chinese health administration.

Our study showed that 10.1% of HCWs enrolled were not willing to participate in the COVID-19 treatment tasks, and the risk of abnormal mental state of these respondents was 2.3 times higher than that of voluntary ones. There was a higher risk of psychological abnormality among HCWs who had been or were being isolated or had family members or colleagues infected. Familial clusters of pneumonia associated with COVID-19 were also observed in and out of Wuhan.^2^ More than half of the respondents expressed concerns about their family and colleagues in risk perception questionnaire. Impressively high contagion rates, long incubation periods and numerous asymptomatic virus carriers^18 19^ may mainly contribute to the HCWs’ concern over their family members, since doctors can’t distinguish the COVID-19 infector in their common clinical work. Some communities even prohibited HCWs from entering the community after work, believing that they may be the potential source of infection, which further aggravated the remorse of HCWs and their concern for their families. As a result, most HCWs have to separate from their families, which further weakens the support from their families. Nevertheless, 89.9% of HCWs voluntarily participated in the frontline treatment of COVID-19.

Only 34.7% of HCWs expressed in the risk perception questionnaire they were very worried about the risk of self-infection. Interestingly, it seems that there is no significant difference in psychological state between respondents with or without self-infection, which is quite unexpected for us. Previous studies of SARS have shown that supports from colleagues, clear instructions and strict precautions helped HCWs cope better with SARS and that they were less likely to develop psychological abnormalities.^20^ A number of studies have suggested that when medical workers are overburdened, their feeling too much responsibility for work, facing the uncertainty of work and ambiguity of role, short of training, and lack of support from superiors and colleagues, are all important exogenous factors causing occupational stresses on medical workers.^21^ The sudden outbreak of COVID-19 resulted in a large number of patients in need of admission and treatment; other professional doctors were also engaged in infectious disease work; medical facilities were incomplete; materials were in short supply; management processes were out of order; and medical personnel were in great shortage due to colleagues’ infection and quarantine--all of these cases aggravated the occupational tension. However, in the context of epidemic disease such as SARS and COVID-19, HCWs fell into a state of unstainable work that they could not give up, resulting in the moral heroism and altruism of the medical group ^22^, as well as the extreme sacrifice of the group, which led to the decline of HCWs’ attention to themselves.

### LIMITATIONS

This study has several limitations. First, because of the disease outbreak, we were unable to conduct face-to-face interviews; second, the results are limited by the use of convenience sampling, which could not reflect the overall status of HCWs in China; and third, due to the time constraints, the scale we used is so simple that it can only provide a preliminary screening, further confirmation and intervention are needed.

## CONCLUSION

In the early stage of COVID-19 epidemic, HCWs, especially in Wuhan, were worried about the risk of infection and protective measures, causing 39.1% of them to have developed psychological distress.

The prevention and control of epidemics should not rely on the sacrifice of a single group. In the subsequent fight against COVID-19 epidemic, adequate protective materials, enough rests, standard operations, protection training for non-infectious medical professionals and strengthening the family support for HCWs are urgent issues. Until this paper was submitted, the novel coronavirus has spread to 73 countries on all continents except Antarctic, European Union raised coronavirus risk level to high from moderate. HCWs in other countries may be confronted with similar difficulties as those in China, and we hoped that their demands and psychological state would be concerned early.

## Data Availability

Individual participant data that underline the results reported in this article, after deidentification will be shared, the data are available form for individual participant data meta-analysis. And the study protocol is available.

## Contributors

Yuhong Dai is first author. Xianglin Yuan and Hong Qiu are joint corresponding authors. Xianglin Yuan and Hong Qiu designed the study. Yuhong Dai, Guangyuan Hu and Huihua Xiong collected the data. Yuhong Dai was involved in data processing. Yuhong Dai drafted the manuscript. Xianglin Yuan and Hong Qiu contributed to the interpretation of the results and critical revision of the manuscript for important intellectual content and approved the final version of the manuscript. All authors have read and approved the final manuscript. Xianglin Yuan and Hong Qiu are the study guarantors.

## Ethical approval

The study was approved by the Ethics Committee of Tongji Hospital of Tongji Medical College of Huazhong University of Science and Technology, and was registered with ClinicalTrials.gov, number NCT04260308.

## REFERENCE

1. Phan LT, Nguyen TV, Luong QC, et al. Importation and Human-to-Human Transmission of a Novel Coronavirus in Vietnam. The New England journal of medicine 2020 doi: 10.1056/NEJMc2001272

2. Chan JF, Yuan S, Kok KH, et al. A familial cluster of pneumonia associated with the 2019 novel coronavirus indicating person-to-person transmission: a study of a family cluster. Lancet 2020 doi: 10.1016/S0140-6736(20)30154-9

3. Wang D, Hu B, Hu C, et al. Clinical Characteristics of 138 Hospitalized Patients With 2019 Novel Coronavirus-Infected Pneumonia in Wuhan, China. Jama 2020 doi: 10.1001/jama.2020.1585

4. National Health Commission of the People’s Republic of China. Report of novel coronavirus-infected pneumonia in China. Published February 4, 2020 (accessed Feb 19, 2020).

5. Nickell LA, Crighton EJ, Tracy CS, et al. Psychosocial effects of SARS on hospital staff: survey of a large tertiary care institution. CMAJ : Canadian Medical Association journal = journal de l’Association medicale canadienne 2004;170(5):793–8. doi: 10.1503/cmaj.1031077

6. Lee AM, Wong JG, McAlonan GM, et al. Stress and psychological distress among SARS survivors 1 year after the outbreak. Canadian journal of psychiatry Revue canadienne de psychiatrie 2007;52(4):233–40. doi: 10.1177/070674370705200405

7. Zhou X, Song H, Hu M, et al. Risk factors of severity of post-traumatic stress disorder among survivors with physical disabilities one year after the Wenchuan earthquake. Psychiatry research 2015;228(3):468–74. doi: 10.1016/j.psychres.2015.05.062

8. Zhou X, Kang L, Sun X, et al. Prevalence and risk factors of post-traumatic stress disorder among adult survivors six months after the Wenchuan earthquake. Comprehensive psychiatry 2013;54(5):493–9. doi: 10.1016/j.comppsych.2012.12.010

9. Cheng TA, Williams P. The design and development of a screening questionnaire (CHQ) for use in community studies of mental disorders in Taiwan. Psychological medicine 1986;16(2):415–22. doi: 10.1017/s0033291700009247

10. Wuhan Municipal Health Commission. Use of designated hospital beds in Wuhan. Published February 4, 2020. (accessed Feb 19, 2020).

11. Zhu N, Zhang D, Wang W, et al. A Novel Coronavirus from Patients with Pneumonia in China, 2019. The New England journal of medicine 2020 doi: 10.1056/NEJMoa2001017

12. WHO. Summary of probable SARS cases with onset of illness from 1 November 2002 to 31 July 2003. Dec 31, 2003. https://www.who.int/csr/sars/country/table2004_04_21/en/ (accessed Feb 19, 2020).

13. The L. Protecting Chinese doctors. Lancet 2020;395(10218):90. doi: 10.1016/S0140-6736(20)30003-9

14. Wang JL YW, Zhang HL, Wang XC. Study on the general mental health status and influencing factors of Chinese Medical Team members. Zhonghua Liu Xing Bing Xue Za Zhi 2019;40(5):5.

15. Firth-Cozens J. Emotional distress in junior house officers. British medical journal 1987;295(6597):533–6. doi: 10.1136/bmj.295.6597.533

16. Ramirez AJ, Graham J, Richards MA, et al. Burnout and psychiatric disorder among cancer clinicians. British journal of cancer 1995;71(6):1263–9. doi: 10.1038/bjc.1995.244

17. Tam CW, Pang EP, Lam LC, et al. Severe acute respiratory syndrome (SARS) in Hong Kong in 2003: stress and psychological impact among frontline healthcare workers. Psychological medicine 2004;34(7):1197–204. doi: 10.1017/s0033291704002247

18. Rothe C, Schunk M, Sothmann P, et al. Transmission of 2019-nCoV Infection from an Asymptomatic Contact in Germany. The New England journal of medicine 2020 doi: 10.1056/NEJMc2001468

19. Guan WJ, Ni ZY, Hu Y, et al. Clinical Characteristics of Coronavirus Disease 2019 in China. The New England journal of medicine 2020 doi: 10.1056/NEJMoa2002032

20. Chan AO, Huak CY. Psychological impact of the 2003 severe acute respiratory syndrome outbreak on health care workers in a medium size regional general hospital in Singapore. Occupational medicine 2004;54(3):190–6. doi: 10.1093/occmed/kqh027

21. Domagala A, Bala MM, Storman D, et al. Factors Associated with Satisfaction of Hospital Physicians: A Systematic Review on European Data. International journal of environmental research and public health 2018;15(11) doi: 10.3390/ijerph15112546

22. Reid L. Diminishing returns? Risk and the duty to care in the SARS epidemics. Bioethic 2005;19(4):348–61. doi: 10.1111/j.1467-8519.2005.00448.x

